# Identification and molecular characterization of missense mutations in orphan GPCR GPR61 occurring in severe obesity

**DOI:** 10.1101/2024.10.21.24315834

**Authors:** Choi Har Tsang, Alexander De Rosa, Paweł Kozielewicz

**Affiliations:** Molecular Pharmacology of GPCRs, Department of Physiology and Pharmacology, Karolinska Institutet, 171 65 Solna, Sweden; School of Engineering Sciences (SCI), KTH Royal Institute of Technology, 114 28 Stockholm, Sweden

## Abstract

Obesity is a disease defined as “abnormal or excessive fat accumulation that presents a health risk, with a body mass index (BMI) over 30” (World Health Organization). Signalling mediated by hypothalamus-expressed membrane proteins from the GPCR family regulates food intake and metabolism. The orphan GPCR GPR61 was linked to regulation of metabolism and, here, we identify 34 mutations in the GPR61 gene which are present with much higher frequency in severe obesity samples from UK10K obesity screen than in the normal population. The cumulative sum of mutations was higher for the GPR61 gene compared to the highly mutated and well-established target, melanocortin 4 receptor (MC_4_R). The majority of the GPR61 mutations reduced cell surface expression of overexpressed HiBiT-GPR61 in HEK293A cells. Additionally, the mutations T92P, R236C and R262C reduced ligand-independent GPR61-induced cAMP production, and R236C compromised G_s_ protein activation. Therefore, we suggest that GPR61 indeed represents a G_s_-coupled receptor with a potential role in the regulation of metabolism. Our data warrant further studies to assess role of this orphan GPCR in metabolism in greater detail.

## Introduction

Obesity is a disease defined as “abnormal or excessive fat accumulation that presents a health risk, with a body mass index (BMI) over 30” (World Health Organization). It is linked to several metabolic problems, including type 2 diabetes and cardiovascular diseases, which contribute to significant morbidity, mortality and increased healthcare costs ^1^. Obesity is associated with abnormal regulation of appetite ^2,3^, and in the human body, the central control of food intake takes place in the hypothalamus ^2,3^. As such, signalling mediated by hypothalamus-expressed membrane proteins from the GPCR family has been shown to regulate food intake and metabolism ^4–8^. Of GPCRs, the proopiomelanocortin (POMC)/melanocortin 4 receptor (MC_4_R) circuit plays a key role in appetite regulation ^6,9,10^. Ligand-induced and constitutive activity of wild-type or gain-of-function mutants of MC_4_R lead to the activation of transducer G_s_ proteins and an increase in intracellular cAMP production, which results in reduced food intake ^6–8^. Furthermore, brain-expressed agouti-related protein (AgRP) acts as an inverse agonist of MC_4_R blocking its signalling ^11^. Interestingly, non-specific G_s_ stimulation in AgRP neurons leads to increased food intake. This suggests that understanding the details of GPCR-G_s_ protein signalling can be key in elucidating molecular mechanisms of the development of obesity ^12^.

GPR61 belongs to the class A orphan GPCRs, i.e. receptors for which endogenous ligands have not been identified and verified by studies from at least two independent laboratories. Here, the incentive to investigate GPR61 was that its role in regulation of metabolism has already been proposed but the receptor is far from being established as a functional regulator of metabolism. Along these lines, GPR61 knockout mice develop significant hyperphagia and increased body weight in comparison to wild-type mice ^13^. In accordance, several genome-wide association studies (GWAS) have predominantly associated reduction in GPR61 expression with increased BMI and body fat composition (https://www.ebi.ac.uk/gwas/genes/GPR61). However only little is known about GPR61-mediated signaling but it is recognized that overexpressed GPR61 constitutively couples to heterotrimeric G_s_, similarly to MC_4_R ^14^. Moreover, and again analogously to MC_4_R, the N-terminus of the receptor plays an important role in its constitutive activity ^15–17^. Furthermore, the structure of GPR61 was resolved by CryoEM using a receptor lacking the N-terminus, the ICL3 and the C-terminus ^18,19^. In these studies, the constitutive activity of the receptor was attributed to the ECL2 penetrating into the putative orthosteric binding pocket. Lastly, weak and potent synthetic inverse agonists have been introduced but the pharmacological toolbox for GPR61 is still relatively limited ^14,18,20^.

Here, we analyzed 480 samples from UK10K obesity datasets containing sequencing information from severe obesity cases (https://www.uk10k.org/studies/obesity.html) and we found 34 missense mutations in the GPR61 gene. Some of these mutations are not present and the rest occur with much higher frequency in diseased individuals than in the normal population (gnomAD). These mutations had impact on cell surface trafficking and constitutive activity of GPR61. Specifically, the R236C mutation led to a significant decrease in constitutve activity of GPR61 in FRET-based cAMP and ebBRET-based G_s_ translocation assays. Furthermore, molecular dynamic (MD) simulations reveal distinct conformational changes in the transmembrane domain structure of the mutated receptor in the comparison with the wild-type counterpart.

In conclusion, this study contributes to the growing body of data that GPR61 presents another G_s_-coupled receptor with a potential role in regulating metabolism. As such, future studies should address the role of this receptor and its mutations in more detail, and in relevant models.

## Materials and methods

### Database search and analysis

We leveraged the UK10K dataset, accessed via the European Genome-Phenome Archive (EGA; dataset IDs: EGAD00001000429, EGAD00001000431, EGAD00001000432; application agreement 15267), to perform comprehensive mutational analysis and single nucleotide polymorphism (SNP) calling. This dataset was selected for its broad genomic coverage, generated using the Illumina HiSeq 2000 platform, and its relevance to the cohort under study, which included individuals with a body mass index (BMI) ≥ 40 or participants in the Severe Childhood Onset Obesity Project (SCOOP). A total of 480 samples (308 from The Severe Childhood Onset Obesity Project (SCOOP), 64 from the TwinsUK study and 108 from The Generation Scotland: Scottish Family Health Study) were available for download and subsequent processing. To ensure data security and integrity, all samples were securely downloaded using the EGA Live Outbox tool. Following this, a series of stringent preprocessing steps were applied to each data sample. The processed reads were then aligned to the GRCh38 human reference genome using TopHat, ensuring high-quality mapping. Acceptable hits were subsequently processed using Samtools mpileup, which provided the foundation for accurate variant detection. For SNP calling and variant detection, we utilized VarScan mpileup, with downstream results converted into interpretable formats using SNPsift. This analytical pipeline ensured precise identification of genetic variants, providing a reliable framework for subsequent analyses. Identified mutations were then cross-referenced with the NCBI Short Genetic Variations database (dbSNP) for validation. Additionally, the Ensembl Variant Effect Predictor (VEP) was employed to predict the functional consequences of each mutation. This tool facilitated the assessment of the potential impact on protein-coding regions, regulatory elements, and associations with known phenotypes, allowing for an in-depth understanding of the genetic variations in this cohort. As controls representing normal population, we used variants from over 730,000 individuals from Genome Aggregration Database (gnomAD v4.1.0; www.gnomad.broadinstitute.org).

### In vitro cell culture

HEK293A cells (Thermo Fisher Scientific) were cultured in DMEM supplemented with 10% FBS (Sigma), 1% penicillin/streptomycin, 1% L-glutamine (both from Thermo Fisher Scientific) in a humidified CO_2_ incubator at 37°C. All cell culture plastics were from Sarstedt, unless otherwise specified. The absence of mycoplasma contamination was routinely confirmed by PCR using 5′-GGCGAATGGGTGAGTAACACG-3′ and 5′-CGGATAACGCTTGCGACTATG-3′ primers detecting 16S ribosomal RNA of mycoplasma in the media after 2–3 days of cell exposure.

### DNA constructs, cloning and mutagenesis

HiBiT-GPR61 plasmid DNA was generated with Gibson cloning using GPR61-Tango plasmid DNA (#66366 Addgene, deposited by Bryan Roth) and HiBiT-FZD_6_ plasmid DNA ^21^ as templates. Plasmid DNA constructs encoding different HiBiT-GPR61 mutants were generated with a GeneArt Site Directed mutagenesis kit (Thermo Fisher Scientific). rGFP-CAAX plasmid DNA and Gαs-67-RlucII plasmid DNA in the pcDNA3.1(+) backbone were synthesized by GeneScript. EPAC-based cAMP sensor H187 plasmid DNA ^22^ was a kind gift from Kees Jalink (The Netherlands Cancer Institute). The constructs were validated by Sanger sequencing (Eurofins GATC).

### Cell surface expression

HEK293A cells were transiently transfected in suspension using Lipofectamine® 2000 (Thermo Fisher Scientific). A total of ca. 4 × 10^5^ cells were transfected in 1 ml with 200 ng of HiBiT-GPR61 WT or mutants plasmid DNA and 800 ng of pcDNA3.1 plasmid DNA, or only 1000 ng of pcDNA3.1 plasmid DNA. Next, transfected cells (4 × 10^4^ cells in 100 μl) were seeded onto white 96-well cell culture plates. Twenty-four hours later, the cells were washed once with 200 µl of HBSS (HyClone). Next, 90 μl of HBSS was added to the wells and subsequently, 10 μl of a mix of furimazine (1:10 dilution; #N2421, Promega) and LgBiT (1:20 dilution; #N2421, Promega) were added. The plate was incubated for 10 min and the NanoBiT emission (460–500 nm, 200 ms integration time) was measured using a Spark microplate reader (TECAN).

### cAMP production

HEK293A cells were transiently transfected in suspension using Lipofectamine® 2000 (Thermo Fisher Scientific). A total of ca. 4 × 10^5^ cells were transfected in 1 ml with 200 ng of HiBiT-GPR61 constructs, 100 ng EPAC-based cAMP sensor and 700 ng of pcDNA3.1 plasmid DNA. Next, transfected cells (4 × 10^4^ cells in 100 μl) were seeded onto black 96-well cell culture plates. Twenty-four hours later, the cells were washed once with 200 µl of HBSS (HyClone). Next, 100 μl of HBSS was added to the wells and FRET ratio measured with a CLARIOstar plate reader (BMG). mTurquoise2 (donor) was excited at 410-450 nm and its emission intensity was measured using a 460-500 nm monochromator. cpVenus173 (acceptor) emission was recorded using a 515-555 nm monochromator. FRET ratios were defined as acceptor emission/donor emission.

### Gα_s_ translocation

HEK293A cells were transiently transfected in suspension using polyethylenimine (PEI, Polysciences). A total of ca. 4 × 10^5^ cells were transfected in 1 ml with 200 ng of HiBiT-GPR61 DNA constructs, 300 ng of rGFP-CAAX plasmid DNA, 40 ng of Gα_s_-67-RlucII plasmid DNA and 460 ng of pcDNA3.1 plasmid DNA. Next, transfected cells (4 × 10^4^ cells in 100 μl) were seeded onto white 96-well cell culture plates. Twenty-four hours later, the cells were washed once with 200 µl of HBSS (HyClone). Next, 90 μl of HBSS was added to the wells and subsequently, 10 μl of coelenterazine 400a, (2.5 μM final concentration, Biosynth). The plate was incubated for 10 min. Next, RlucII emission (donor, 360-440 nm, 200 ms integration time) and rGFP emission (acceptor, 505-575 nm, 200 ms integration time) were measured using Spark microplate reader (TECAN). The BRET2 ratios were defined as acceptor emission/donor emission.

### Molecular dynamics simulations

Molecular dynamics (MD) simulations of GPR61 were conducted using a high-resolution structural model (Protein Data Bank ID: 8KGK) as input. The simulations were initiated on the CHARMM-GUI server. The unresolved C-terminal region of GPR61, missing from the Cryo-EM structure, was not modelled and deliberately excluded from the simulations. To mimic the native membrane environment, a hexagonal phospholipid bilayer was carefully constructed around the protein using a lipid composition representative of a physiological membrane. This bilayer was embedded in a solution of water molecules and 0.15 M NaCl to replicate typical ionic conditions in biological systems. The CHARMM36m force field was employed to model the molecular interactions, ensuring that all protein-lipid and protein-solvent interactions were treated with high accuracy. The protonation states of the residues were assigned at a physiological pH of 7.4, optimising the protein’s charge states for biological relevance. Energy minimisation was performed on the system using the steepest descent algorithm to eliminate any steric clashes and relieve strain, followed by a multi-step equilibration process. Furthermore, during the production phase, the simulations was conducted under NPT conditions, ensuring constant pressure and temperature throughout. The system’s temperature was regulated at 310 K using the Nose–Hoover thermostat. Each mutational variant and wild type of GPR61 underwent triplicate simulations, with each trajectory spanning 200 ns, leading to a total simulation time of 600 ns per variant. Upon completion, all MD trajectories were analyzed using Chimera (version 1.17.3) or VMD. RMSDs from individual runs can be found in the **Supplementary Figure 1**.

### Statitstical analysis

All data presented in this study come from at least three individual experiments (biological replicates) with each individual experiment performed typically at least in duplicates (technical replicates) for each condition, unless otherwise specified in a figure legend. One biological replicate is defined as wells with cells seeded from the same individual cell culture flasks and measured on the same day. Different biological replicates were transfected using separate transfection mixes. Technical replicates are defined as individual wells with cells from the same biological replicate. Samples were not randomized or blinded during the experiments. Statistical and graphical analyses were performed using Graph Pad Prism software. Data were analyzed by one-way ANOVA with Fisher’s least significant difference (LSD) post-hoc analysis. Significance levels are given as: \**P* < 0.05; \*\**P* < 0.01; \*\*\**P* < 0.001; \*\*\*\**P* < 0.0001. Data points throughout the manuscript are indicated as the mean ± standard error of the mean (s.e.m.)

## Results

### Cumulative sum of GPR61 mutations is higher than for MC_4_R in the analysed severe obesity samples

In our comprehensive analysis of 480 severe obesity samples (**Figure 1**), we identified 34 different missense mutations in the GPR61 gene (**Figure 2A**). Multiple mutations appeared more than once with the V287L being the mutation, which occurred with the highest frequency: in nine samples. The cumulative sum of GPR61 mutations was 73. Importantly, the following nine mutations: T92P, H146R, V152A, G165V, L215F, L218P, L223V, Q273E and L289F were absent from the gnomAD database (**Supplementary Table 1**). The mutations are localized throughout the polypeptide chain excluding the C-tail (**Figure 2B**). Except for S122, the amino acids at these positions are not predicted to be involved in stabilization of inactive or active state of GPCRs (https://gpcrdb.org/structure_comparison/comparative_analysis#) ^23^. However, many identified mutations in GPR61 are predicted to have a negative impact on the structure and function of the receptor (**Supplementary Figure 2**) ^24^. Next, we extended our analysis to two additional genes MC_4_R and GPR141. As mentioned, MC_4_R is implicated in metabolic regulation, and presents with a very high mutational rate in obese samples ^25–27^. We have identified 50 missense mutation variants of MC_4_R in the cohort (**Supplementary Figure 3A**) including previously undescribed mutations. The MC_4_R mutations occurred only one time each. In contrast, 13 mutations were identified in the GPR141 gene (**Supplementary Figure 3B**). GPR141 is a GPCR with no known direct association with obesity ^28^.

**Figure 1.**
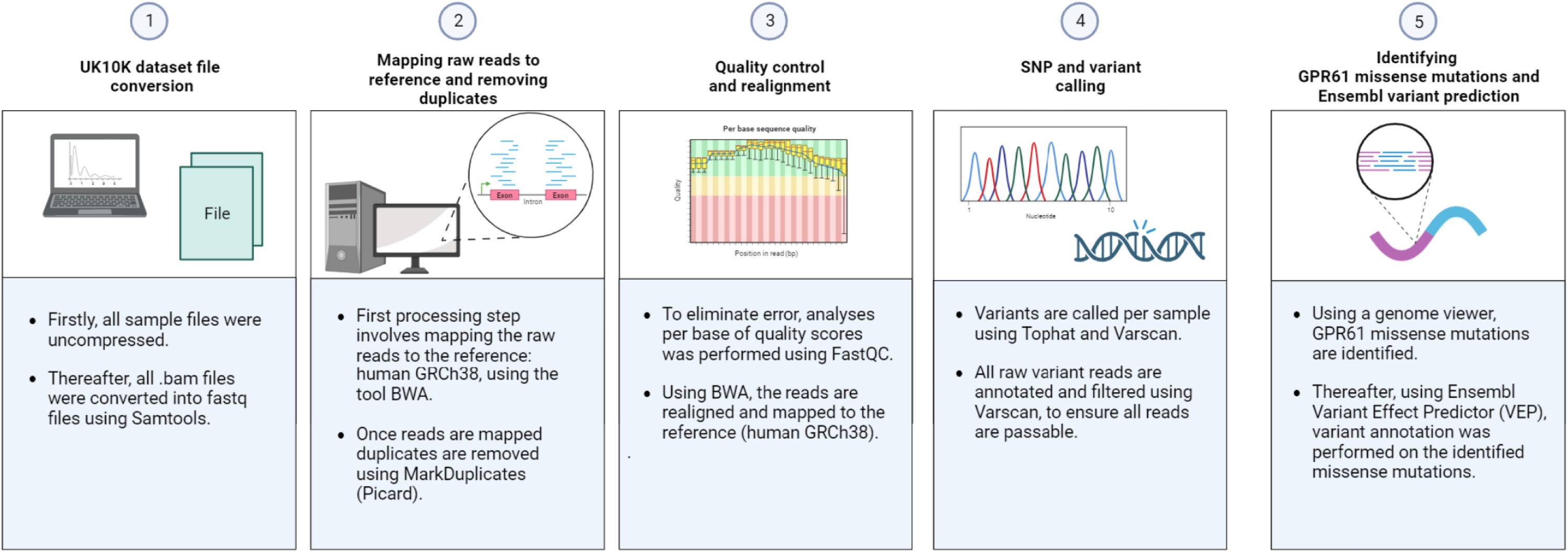
Protocol for exon sequencing and SNP variant calling. The same procedure was used for GPR61, MC_4_R and GPR141.

**Figure 2.**
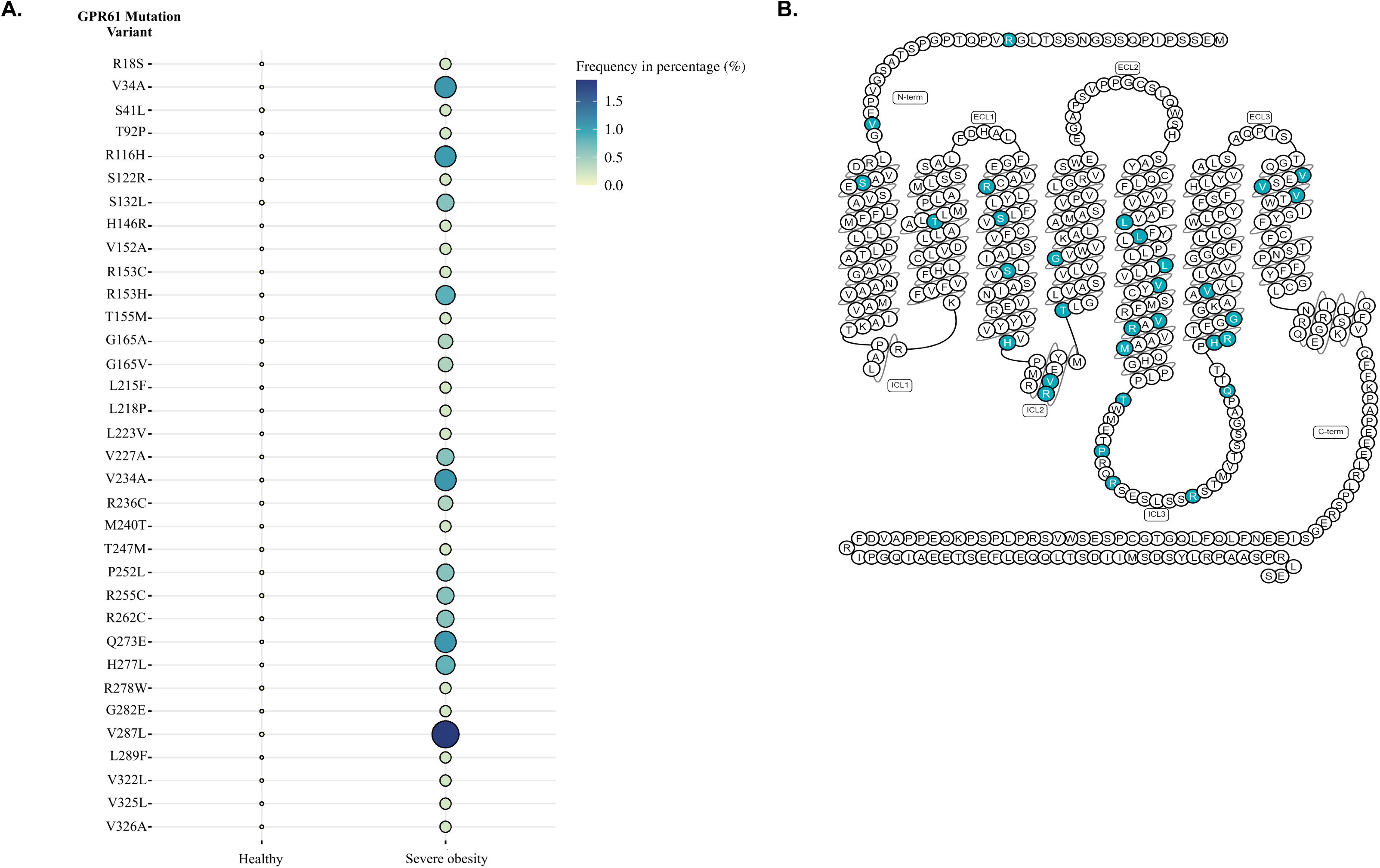
UK10K obesity screen reveals presence of GPR61 mutations. **A.** Mutational landscape of GPR61 in normal population and then the same mutations in severe obesity patients. **B.** GPR61 mutations mapped onto a 2D model of GPR61.

### GPR61 mutations reduce cell surface expression

We have generated 34 different HiBiT-GPR61 plasmid DNA constructs, each carrying a single mutation as found in the UK10K obesity datasets analysis. Using NanoBiT luminescence measurements, we have analyzed cell surface expression of the constructs upon overexpression in HEK293A cells. We could demonstrate that 18 mutations lead to a statistically-significant reduction of cell surface expression and two mutations, S132L and G165V, lead to a statistically-significant increase of receptor cell surface expression (**Figure 3**).

**Figure 3.**
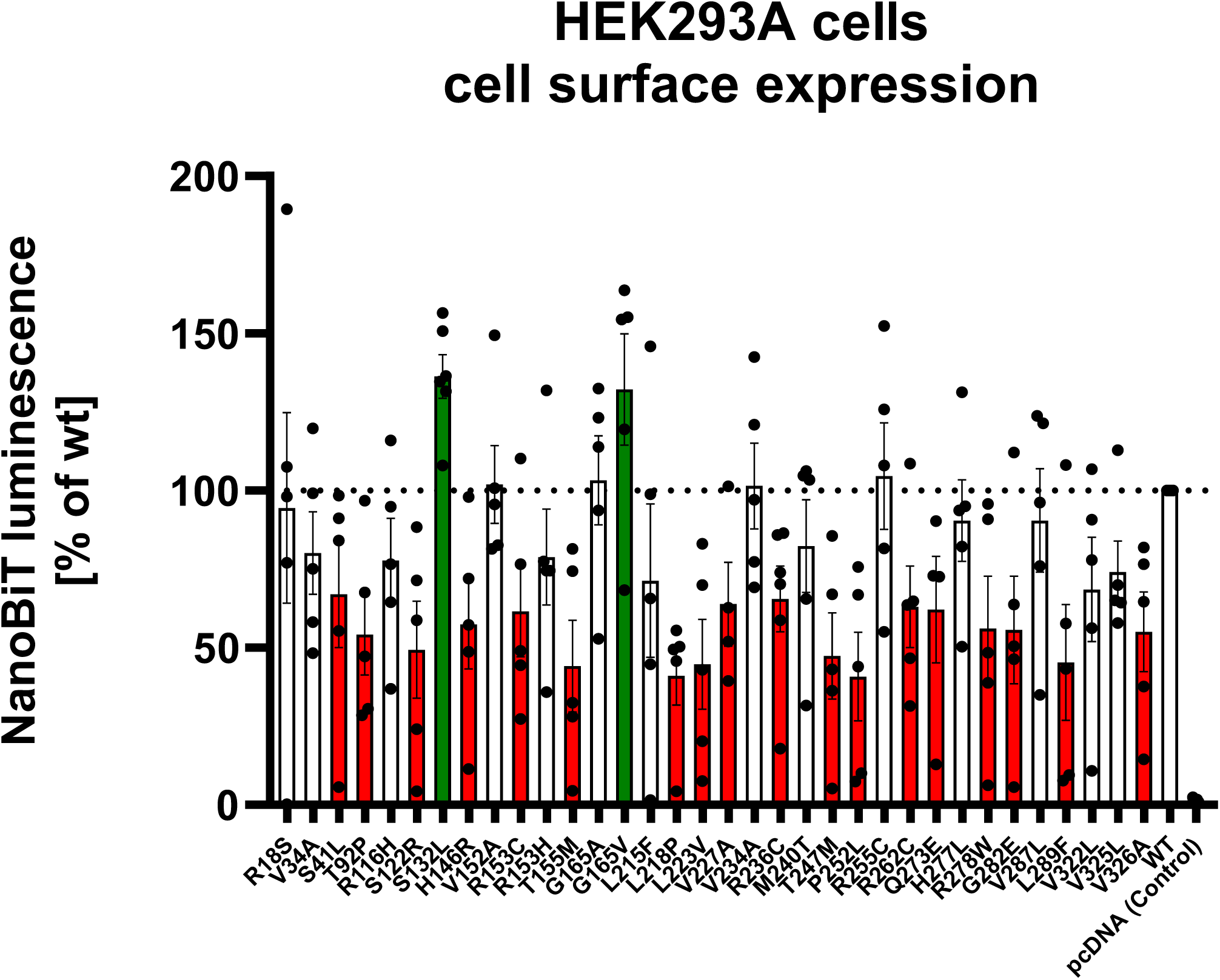
Obesity-associated mutations of GPR61 have impact on cell surface expression. NanoBiT-based measurement of cell surface expression of 34 mutants of GPR61. The red bars indicate statistically significant decrease and the green bars indicate statistically significant increase in NanoBiT-emmitted cell membrane-associated luminescence indicative of cell surface expression of HiBiT-tagged GPR61 constructs; at least p<0.05 and at least n=3 independent experiments.

### GPR61 mutations reduce constitutive activity of the receptor

Using a genetically encoded, EPAC-derived FRET-based cAMP biosensor, we could show that three mutations (T92P, R236C and R262C) increased the FRET ratio of the probe in comparison to the wild-type receptor suggesting inhibition of constitutive GPR61-induced production of cAMP (**Figure 4A**). Next, we focused on these three mutations and performed an orthogonal ebBRET-based G_s_ translocation assay as a measure of heterotrimeric G_s_ activation ^29^. The results from these experiments mirrored the data from cAMP production experiments only for the GPR61 R236C (**Figure 4B**).

**Figure 4.**
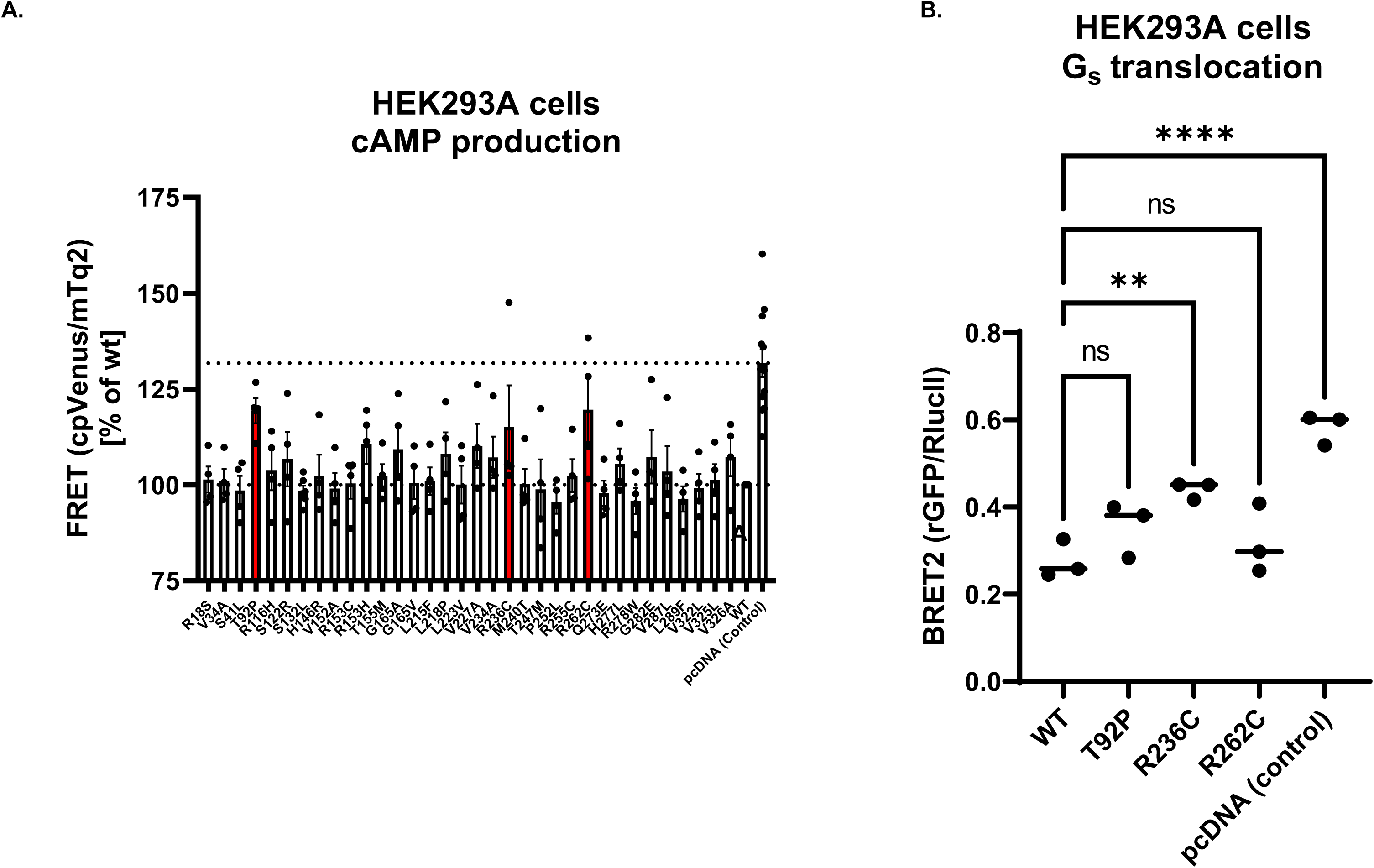
Obesity-associated mutations of GPR61 have impact on cAMP production and G_s_ translocation. **A.** EPAC-derived FRET-based biosensor reveals that three mutations (the red bars) lead to the statistically significant reduction in cAMP production elicited by overexpression of a GPR61 in the absence of an agonist (increase in the FRET ratio reflects decrease in cAMP levels). **B.** The presence of overexpressed GPR61 R236C leads to a statistically-significant reduction in the bystander BRET ratio between Gαs-67-RlucII and rGFP-CAAX indicative of reduced G_s_ translocation away from the cell membrane; at least p<0.05 and at least n=3 independent experiments.

### MD simulations reveal changes in receptor structure for GPR61 R236C

In our molecular dynamics (MD) simulations of the R236C mutation, chosen based on *in vitro* data from the constitutive activity studies, we observed significant structural deviations compared to the wild-type protein. Structurally, the GPR61 wild-type at position 236 has an arginine that presents itself with a helix. The mutatnt contains a cysteine residue which resulted in a loss of alpha helical structure, and presence of different intrahelical interactions (**Figure 5**). To this end, in the wild-type R236 forms Van der Waals, hydrophobic and polar interactions with F232, polar with R233, polar with A235, polar with V237 and polar with A238. In the mutant, C236 forms polar and Van der Waals interactions with F232, polar interactions with R233 and A235.

**Figure 5.**
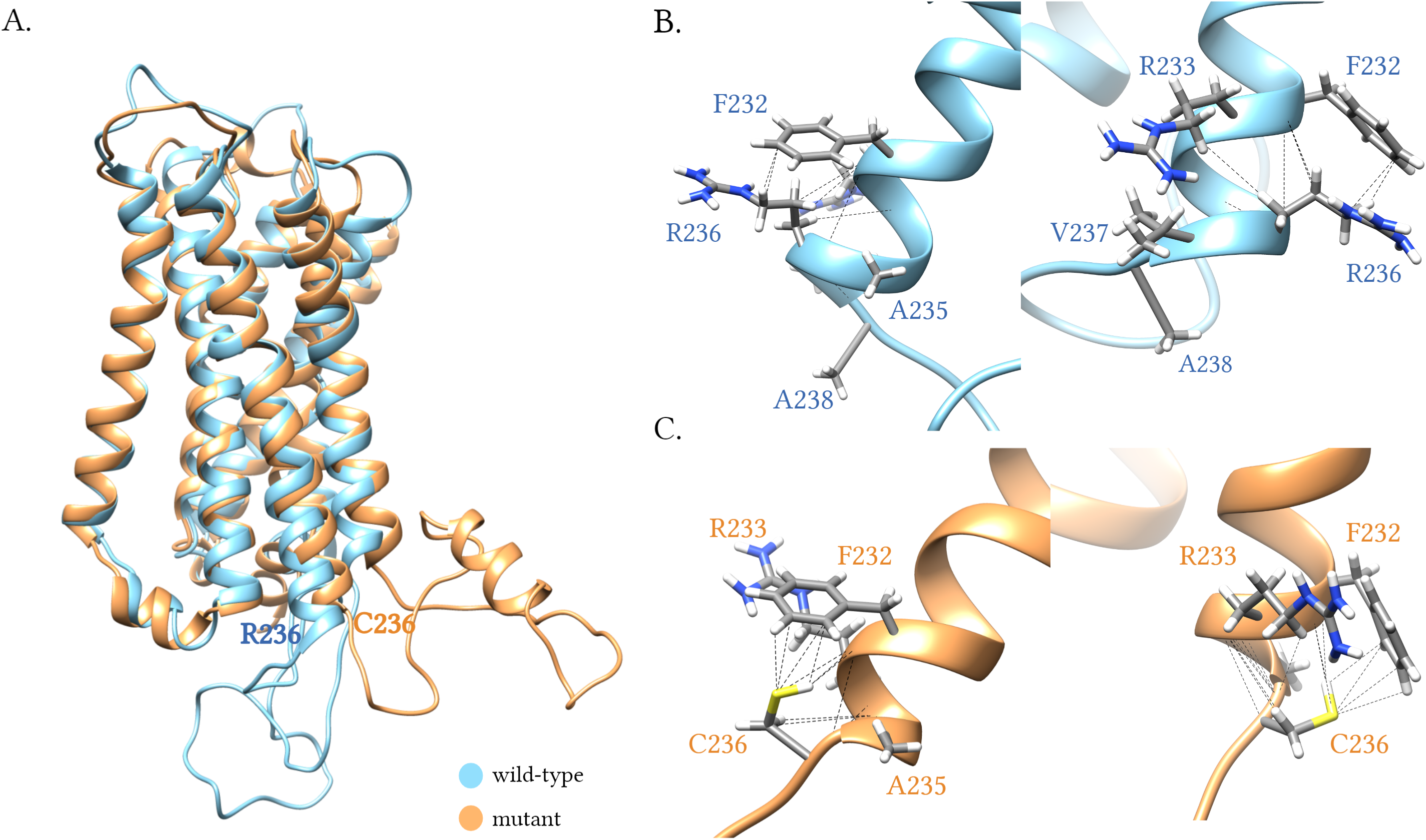
Molecular dynamics simulations predict changes in the structure of the helix 5 following introduction of the R236C mutation. **A.** Final receptor conformations after 200 ns simulation from one representative MD simulation. **B.** Zoomed-in region of R236 (wild-type) and **C.** C236 (mutant) with polar interactions depicted as dashed lines. RMSDs from the independent runs can be in the **Supplementary Figure 1**.

## Discussion

The hypothesis of this study was that GPR61 is important for the regulation of metabolism, and that this role would be reflected by a relatively high number of mutations present in samples from severily obese individuals. This hypothesis was tested mainly through drawing analogies between GPR61 and MC_4_R. MC_4_R, is a well-characterized receptor involved in regulating appetite and its variants are the key contributors to monogenic obesity, affecting satiety signals and homeostasis energy expenditure ^8,10,25,27,30,31^. Another incentive for this study was to test the notion emerging from a few previously published studies that an orphan GPCR GPR61 has a role in metabolic regulation and if so, postulate that it can be grouped with other GPCRs with established links to obesity (e.g. GLP-1, GPR35). The study was not aimed to assess or to prove causation between presence of GPR61 mutations and (severe) obesity. It has been shown that obese individuals have increased rate of DNA instability, elevated number of mutations and dysregulation in cAMP/PKA signaling axis ^32–34^.

First, we have analyzed samples from UK10K obesity screen for the presence of missense mutations in GPR61 gene and compared them with the normal population found in the gnomAD database. We compared the frequency of mutations in GPR61 with two other GPCRs: MC_4_R and GPR141 to assess whether similar patterns of mutation frequency could be observed in genes with established roles (MC_4_R) and uknown roles (GPR141) in obesity and metabolic disorders. The presence of these mutations provided a comparative baseline for the GPR61 findings, given MC_4_R’s established role in obesity. There are no studies linking GPR141 function to metabolism ^28^. Collectively, the analyses of MC_4_R, and GPR141 provided a suitable framework to support a link between the presence of GPR61 mutations identified in samples from individuals and severe obesity. We use this analogy to postulate that the high frequency of mutations observed in GPR61, and even a higher cumulative number of mutations than for MC_4_R, suggests a functional role of GPR61 in metabolism/obesity. In fact, there were 16 mutations in GPR61 which occurred more than once in our analysis, whereas each of identified MC_4_R mutations appeared only once. Along these lines, the limited number of mutations detected in GPR141, a gene unrelated to obesity, together with their low occurrence frequency, further supports our aforementioned hypothesis about the relevance of the GPR61 findings in the context of obesity. Moreover, the absence of some of the mutations in gnomAD underlines that they have not yet been documented in the broader genetic landscape.

Next, we assessed the impact of the mutations on cell-membrane expression and constitutive activation of the receptor using heterologously-overexpressed receptor plasmids in HEK293A cells. The receptor constructs used in this study are N-terminally tagged with HiBiT, which enables selective quantification of cell surface expression following addition of cell-impermeable LgBiT and subsequent measurements of cell surface specific NanoBiT-associated luminescence. We demonstrated that, more than 50% of the identified mutations affected membrane trafficking of the receptor in comparison with the wild type GPR61, predominantly by reducing cell surface expression. Subsequently, we assessed the impact of the mutations on the receptor activation profile. There is no validated agonist for GPR61 available and therefore we could not assess its ligand-mediated activity, but were rather restricted to quantifying its constitutive activity. GPR61 couples to G_s_ and we employed two experimental paradigms to measure G_s_-linked activity of this receptor: EPAC-derived FRET-based biosensor for cAMP and an ebBRET-based G_s_ translocation/activation assay. In the cAMP assay, the mutations T92P, R236C and R262C reduced GPR61-mediated cAMP accumulation. However, in an orthogonal G_s_ translocation assay, it was only R236C that lead to statistically-significant differencs in ebBRET between itself an the wild-type receptor. We hypothesize, that the differences between the results from cAMP and G_s_ activation experiments originate in 1) different signal amplifications of the signaling level and 2) a lack of linear correlation between G_s_ activation and cAMP production. ^35,36^.

Finally, we performed MD experiments to simulate the molecular landscape of the GPR61 carrying C262 in comparison to WT. Our MD simulations on the R236C mutant showed that arginine to cysteine mutation alters the arrangement of helix 5. The residue interaction contacts were different between the two conformations, with the WT establishing five contacts and the mutant having three. This difference in contacts in the mutant can be attributed to the unique chemical properties of cysteine. Cysteine’s thiol group (–SH) is more reactive and capable of forming additional interactions, potentially through increased hydrogen bonding or disulfide bridge formation, leading to greater interaction density. The MD simulations also demonstrate a different orientation of the ICL3 between the mutant and the WT receptor which could potentially intercalate G_s_ coupling. However, the ICL3 had to be modelled in our system and therefore these are only weak predictions. Overall, these structural and interaction differences could explain the mutation’s impact on protein stability and function despite maintaining similar overall contact patterns.

The obvious limitation of this study is that the lack of a GPR61 agonistic ligand substantially restricts our analyses of mutants’ activity. Next, it remains unclear if and how results from studies employing overexpressed receptors in HEK293 cells can be translated to native settings to draw any conclusions on signaling and role of GPR61 in obesity, particularly given that the levels of physiological expression of GPR61 RNA and protein in seemingly the most relevant organ for metabolism and food intake regulation, brain are only moderate (https://www.proteinatlas.org/ENSG00000156097-GPR61/brain). Nevertheless, we conclude that our findings contribute to the general knowledge about GPR61’s role in health and disease, and add relevant information about severe obesity-linked mutations of this receptor in the overexpressed conditions. Our data lay the basis for further studies on the mechanistic understanding of this receptor and its role in obesity in (patho-)physiologically-relevant models.

## Abbreviations

GPCR: G protein-coupled receptor
BRET: bioluminescence resonance energy transfer
FRET: Förster resonance energy transfer
ICL3: intracellular loop 3

## Acknowledgments

PK acknowledges Prof. Nicholas M. Barnes (University of Birmingham, UK) who very effectively convinced me to investigate GPR61 back in 2015. We acknowledge Prof. Gunnar Schulte (Karolinska Institutet) for support. Computations were performed at NSC Tetralith provided by the National Academic Infrastructure for Supercomputing in Sweden (NAISS) funded by the Swedish Research Council through grant agreement no. 2022-06725 (NAISS).

## Data Availability Statement

The authors declare that the majority of the data supporting the findings of this study are contained within the paper. MD data that support the findings of this study will be deposited on gpcrmd.org buty they also are available on request from the corresponding author.

## Authorship Contributions

Participated in research design: Kozielewicz

Conducted experiments: Tsang, De Rosa, Kozielewicz

Contributed new analytic tools: Tsang, Kozielewicz

Performed data analysis: Tsang, De Rosa, Kozielewicz

Wrote or contributed to the writing of the manuscript: Tsang, Kozielewicz

---

**Supplementary Figure 1.**
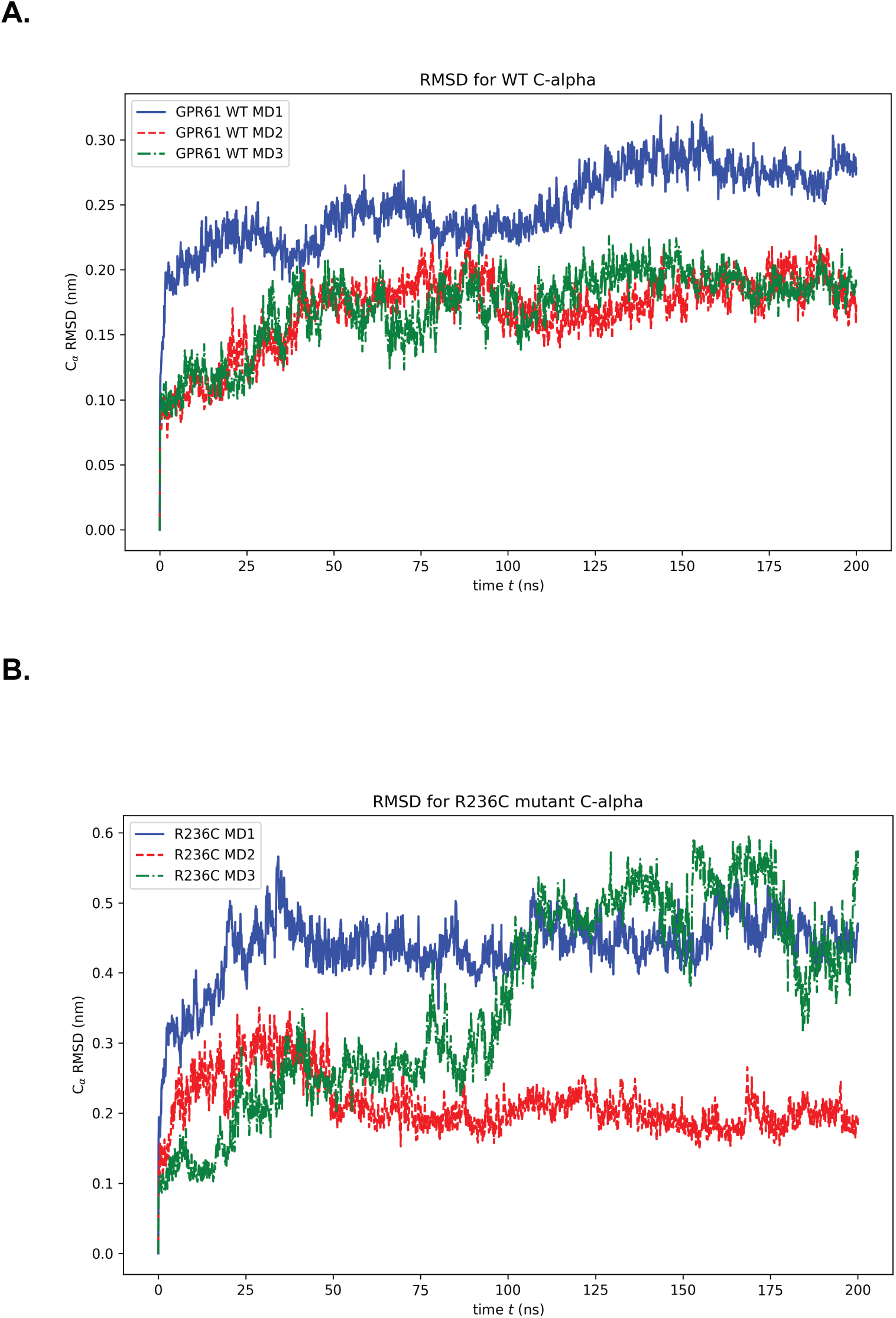
RMSD of carbon alpha atoms from the three simulations of the GPR61 WT (**A**) and GPR61 R236C (**B**).

**Supplementary Figure 2.**
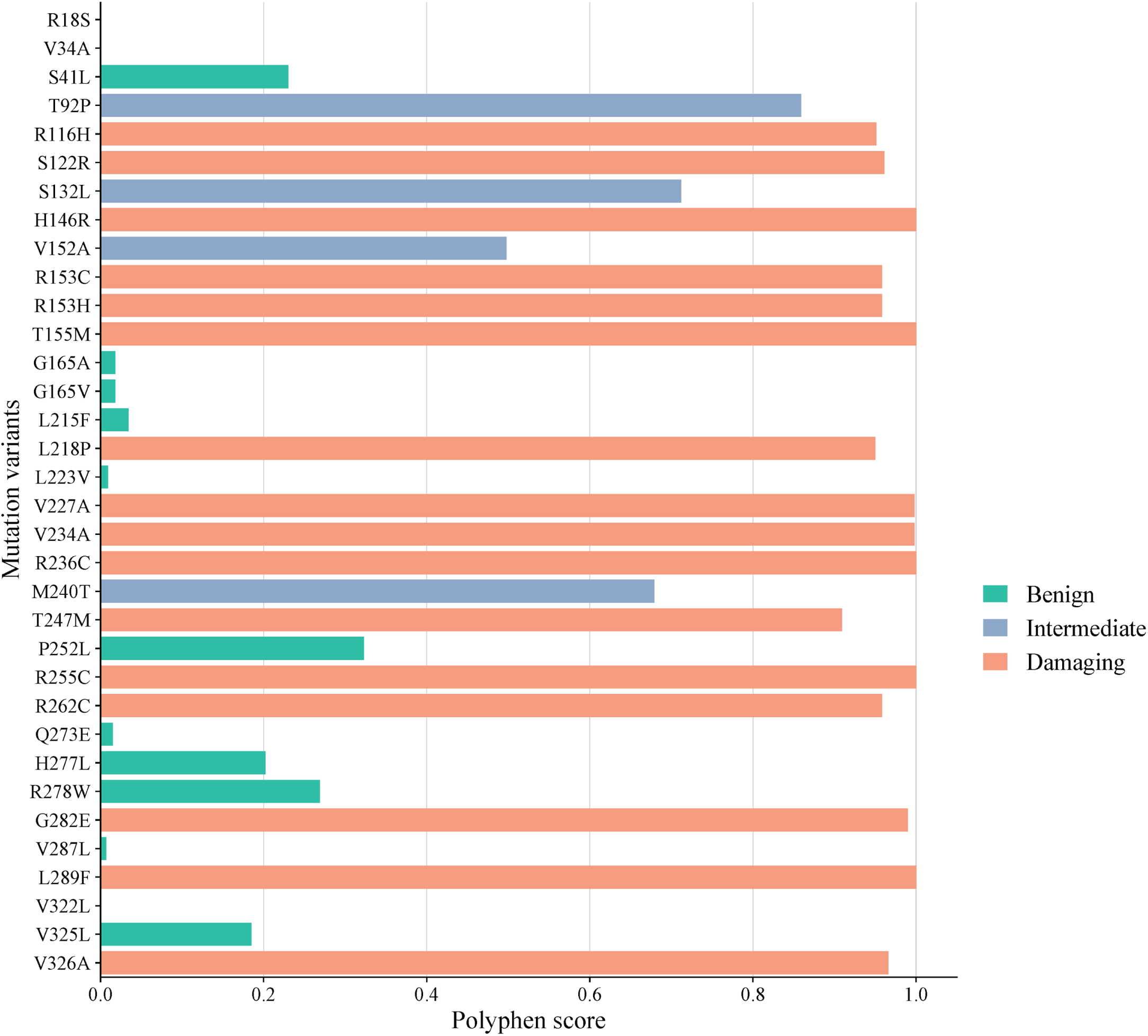
PolyPhen-2 analysis of the 34 mutations in GPR61 to predict their impact on structure and function of the protein.

**Supplementary Figure 3.**
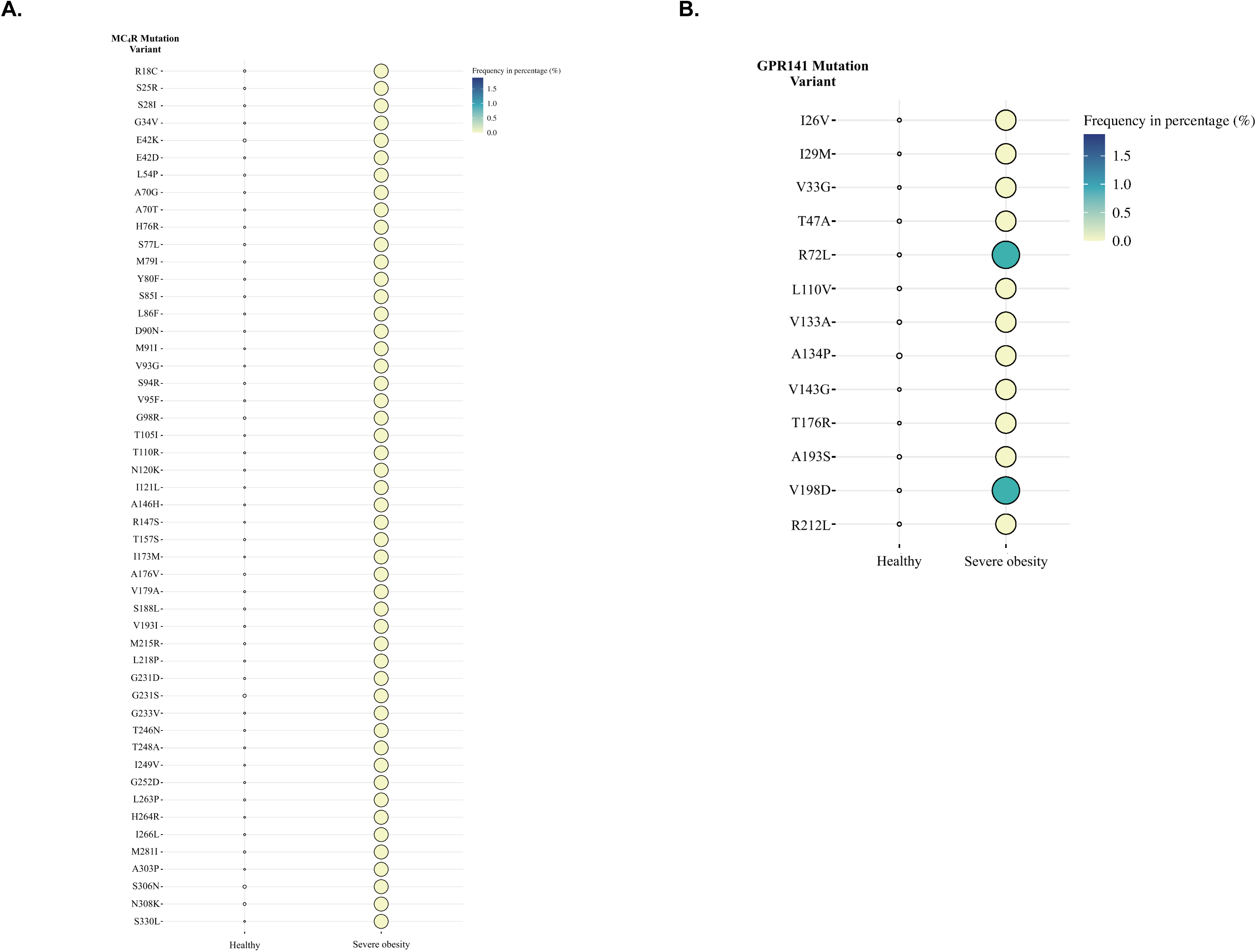
Mutational landscape of MC_4_R (**A**) and GPR141 (**B**) in normal population and then the same mutations in severe obesity patients.

